# “Self-care selfies”: Patient-uploaded videos capture meaningful changes in dexterity over 6 months

**DOI:** 10.1101/2023.08.21.23294268

**Authors:** Arpita Gopal, Wilson O. Torres, Ilana Winawer, Shane Poole, Ayushi Balan, Hannah S. Stuart, Nora E. Fritz, Jeffrey M. Gelfand, Diane D. Allen, Riley Bove

**Affiliations:** University of California, San Francisco Department of Neurology; University of California, Berkeley Department of Mechanical Engineering; University of California, San Francisco Department of Physical Therapy and Rehabilitation Sciences; Wayne State University, Department of Neurology and Program of Physical Therapy

## Abstract

**Objective:** Upper extremity function reflects disease progression in multiple sclerosis (MS). This study evaluated the feasibility, validity and sensitivity to change of remote dexterity assessments applying human pose estimation to patient-uploaded videos.

**Methods:** A discovery cohort of 50 adults with MS recorded “selfie” videos of self-care tasks at home: buttoning, brushing teeth, and eating. Kinematic data were extracted using MediaPipe Hand pose estimation software. Clinical comparison tests were: grip and pinch strength, 9 hole peg test (9HPT), and vibration, and patient-reported dexterity assessments (ABILHAND). Feasibility and acceptability were evaluated (Health-ITUES framework). A validation cohort (N=35) completed 9HPT and videos.

**Results:** The modality was feasible: 88% of the 50 enrolled participants uploaded ≥3 videos, and 74% completed the study. It was also usable: assessments easy to access (95%), platform easy to use (97%), and tasks representative of daily activities (86%). The buttoning task revealed 4 metrics with strong correlations with 9HPT (nondominant: r=0.60-0.69, dominant: r=0.51-0.57, p<0.05) and ABILHAND (r=-0.48, p=0.05). Retest validity at 1 week was stable (r>0.8). Cross-sectional correlations between video metrics and 9HPT were similar at 6 months, and in the validation cohort (nondominant: r=0.46, dominant: r=0.45, p<0.05). Over 6 months, pinch strength (5.8 to 5.0kg/cm^2^, p=0.05) and self-reported pinch (ABILHAND) decreased marginally. While only 15% participants worsened by 20% on 9HPT, 70% worsened in key buttoning video metrics.

**Interpretation:** Patient-uploaded videos represent a novel, patient-centered modality for capturing dexterity that appears valid and sensitive to change, enhancing its potential to be disseminated for neurological disease monitoring and treatment.

## Introduction

The hand is the most active part of the upper extremity, and hand function is critical to independence in activities of daily living (ADLs).^1^ Neurological conditions can result in worsening in hand function, whether acutely (stroke) or more gradually (multiple sclerosis (MS) or Parkinson’s Disease). Changes in function and manual dexterity can affect quality and independence of performance in daily tasks, ability to perform at work and remain employed, and engagement in recreational activities.^2,3^ Assessments sensitive to changes in hand function could inform disease progression, or response to pharmacological^4^ and rehabilitation strategies. To this end, remote assessments have been explored; existing options – smartphone-based applications,^5,6^ wrist-worn accelerometers,^7,8^ and specialized keyboards^9,10^—pose challenges in either implementation, scalability, or relevance to ADLs.

Prioritizing the needs and preferences of patients, patient-uploaded videos represent an entirely different approach to the remote monitoring of dexterity. They leverage accessibility (“bring your own device”, i.e., use of preferred devices and software), usability (video “selfies”: a convenient, modern communication medium), and meaningfulness (capturing self-care tasks critical to independent living). To analyze these videos, pose estimation: a set of algorithms capable of detecting body landmarks and their movement in space, is gaining acceptance in healthy populations (e.g., in the lower extremity to quantify features of gait^11,12^). To date, however, pose estimation has mostly been validated as a method for capturing precise data on movement in healthy human populations, and not to our knowledge in MS or other neurological populations.

The current study sought to validate patient-uploaded “self-care selfies” as a method of remotely monitoring dexterity data for a neurological population. Specific aims were to (1) evaluate the feasibility of video collection at single and longitudinal timepoints, (2) validate dexterity measures extracted from videos using a proof-of-concept, open-source pose estimation algorithm, and (3) evaluate whether subtle worsening in hand function over 6 months could be captured in videos. The overarching goal is to provide a granular, clinically meaningful metric of movement in the natural environment that can be used to measure progression and/or improvement in clinical trials and to inform clinical care and neurological rehabilitation.

## Methods

### Study Setting and Participants

Discovery Cohort. A convenience sample of 50 adult patients with a confirmed diagnosis of MS (by 2017 McDonald Criteria) were referred by their primary neurologist at the University of California, San Francisco (UCSF) Multiple Sclerosis Center to the study team. All referred participants met the additional eligibility criteria (smartphone ownership, smartphone literacy test).

### Study procedures

At baseline and 6 months, in-clinic assessments of hand function were: grip and pinch strength (Jamar Technologies dynanometer)^13^, quantitative vibration sense (Vibratron II-Physitemp)^14^ and the 9HPT (standard for assessing dexterity in MS).^15^ The Action Research Arm Test (ARAT) was also included as a comparsion in-clinic assessment, but was not included in analysis as the cohort reached a ceiling effect. Participants were then trained (for approximately 5 minutes) to use the institutionally approved Research Electronic Data Capture (REDCap) platform ^16^ to complete the remote patient reported outcome (PRO), ABILHAND (23 items summarized in pinch and grip subscores),^17^ and to upload their videos.

#### Video tasks

Participants self-recorded videos of performance of 3 basic ADLs that require hand function^18^: dressing (buttoning a shirt), personal hygiene (brushing teeth), and feeding (fork to mouth). Example videos were provided within REDCap. Feeding and personal hygiene tasks were uploaded once per hand, for a total of 5 videos per timepoint.

#### Study feasibility and acceptability

This was evaluated via monthly REDCap survey using the validated Health Information Technology Usability Evaluation Scale (ITUES)^19^ framework, which evaluates usability to ultimately inform the ease of adoption of new technologies.

### Validation cohort (cross-sectional)

To ensure that the key cross-sectional findings were stable, a second sample of 35 patients meeting the same eligibility criteria was enrolled to provide the following information: baseline demographic and clinical measures, 9HPT and self-care videos.

### Ethical approvals

All study activities were approved by the UCSF IRB (IRB# 20-557) and all participants provided written informed consent.

### Video analysis

Pose estimation has been tested and successfully validated in healthy populations.^20,21^ To estimate 3D hand pose from 2D patient-uploaded videos, the freely available machine learning solution, MediaPipe Hand from the OpenCV library was used. MediaPipe Hand detects 21 landmarks in the hand, including the wrist. Based on the variable grips required for each of the ADLs, the tip of the index finger and wrist landmarks were chosen for analysis (Figure 1). For each landmark, position and velocity kinematic data was obtained by mapping the joints of interest into a xy-cartesian coordinate system (Figure 1). The following metrics were generated: 1) path length of the landmark, a measure of the joint movement in the cartesian plane; 2) complexity of movement, a count of the number of local peaks in the position vs time curve; 3) total distance the joint travels, the area under the velocity vs time curve (AUC); and 4) smoothness, average velocity divided by maximum velocity (Supplementary Table 1).

**Figure 1.**
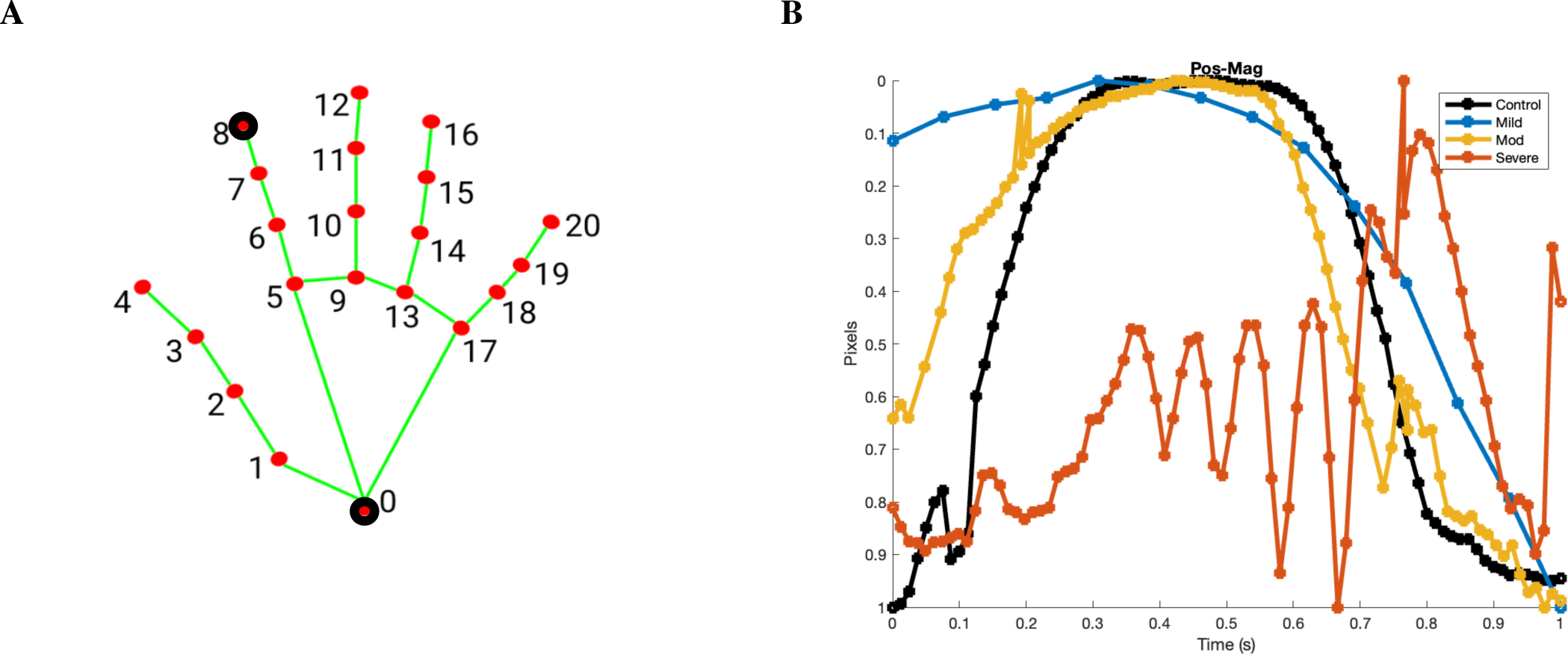
Pose estimation landmarks for video analysis. (A) Depiction of skeleton overlay in MediaPipe Hand. Index finger and wrist landmarks are highlighted in yellow. Coordinate system for video kinematics, with O at the origin. (B) All available landmarks in MediaPipe Hand. Point 8 represents the index finger, and Point 0 represents the wrist. (C) Path length trajectory for index finger in eating task over 1 second. Each participant is depicted by a different colored line (black: control; blue: participant with mild MS (EDSS 0-2.5); yellow: participant with moderate MS (EDSS 3-5.5); red: person with severe MS (EDSS >6). Distance is measured in pixels.

A video was considered valid if ≥10 coordinates were generated during analysis. Sampling frequency was established based on the frame rate of the uploaded video.

As a quality control measure, joint trajectories generated using MediaPipe were validated against Kinovea,^22^ a video annotation software for motion analysis, in 5 randomly selected videos. No statistically significant differences emerged between the manual analysis and MediaPipe (p>0.05).

An initial script was developed to analyze videos using MediaPipe; this is now optimized to automatically generate data on 1 video per minute and available on GitHub.

### Statistical analyses

To describe the study population, feasibility, and correlates of adherence, descriptive statistics and unpaired t-tests were used. To determine the associations between demographic data, video kinematics, PROs, and clinical dexterity measures, Spearman’s correlation coefficients were calculated. To quantify statistically significant changes in each clinical and video measure over the study period, paired t-tests were conducted. Given the large number of variables available through MediaPipe, it was *a priori* decided that the task with the strongest cross-sectional correlations (r > 0.60) with 9HPT would be selected for longitudinal analyses. Spearman’s correlation coefficients were calculated to determine correlations between clinical and video metrics at both 0 and 6 months. Video analyses were performed using Python 3.4; kinematic and statistical analyses, and data visualizations were performed in Matlab 2022.

## Results

### Participant Demographic and Clinical Characteristics

Among the 50 enrolled Discovery cohort participants, 62% were women and 73% non-Hispanic White; mean age was 47.2 years (SD 12.9), and median patient-reported EDSS^23^ disability was 3 (IQR 2, 5) (Table 1). Based on a 9HPT cutoff of 33.3 seconds, 90% of participants had high and 10% had low dexterity.^24^ In-clinic dexterity measures showed some associations with demographic (older age, female sex) and clinical (MS duration) features (Supplementary Figure 1). The Validation cohort did not differ in any key features (p>0.05 for each comparison, Supplementary Table 2).

**Table 1.** Discovery cohort demographics and baseline characteristics.

|  |  |
| --- | --- |
| Gender [N (%)] |  |
| Men | 18 (36%) |
| Women | 31 (62%) |
| Non-binary | 1 (2%) |
| Age [mean (SD)] | 47.2 (12.9) |
| Race [N (%)] |  |
| White, non Hispanic | 44 (73%) |
| Black | 1 (2%) |
| Asian | 4 (8%) |
| Hispanic | 1 (2%) |
| Baseline EDSS [median (IQR)] | 3 (2, 5) |
| Disease duration (years) [mean (SD)] | 14.3 (9.2) |
| MS Subtype [N (%)] |  |
| Relapsing remitting | 43 (86%) |
| Primary progressive | 4 (8%) |
| Secondary progressive | 3 (6%) |
| Dominant hand [N (%)] |  |
| Right | 45 (90%) |
| Left | 5 (10%) |
| Smartphone type [N (%)] |  |
| iOS | 40 (80%) |
| Android | 10 (20%) |
| Grip strength (kg/cm<sup>2</sup>) [mean (SD)] |  |
| Dominant | 28.6 (9.4) |
| Non-dominant | 27 (9.5) |
| Pinch strength (kg/cm<sup>2</sup>) [mean (SD)] |  |
| Dominant | 5.8 (2.6) |
| Non-dominant | 5.6 (2.5) |
| Nine-hole peg test (seconds) [mean (SD)] |  |
| Dominant | 23.5 (9.7) |
| Non-dominant | 28.7 (14.9) |
| Vibration sense (Hertz) [mean (SD)] |  |
| Dominant | 2.4 (2.7) |
| Non-dominant | 2.3 (2.6) |
| ABILHAND [mean (SD)] |  |
| Pinch subscore | 29.5 (4.6) |
| Grip subscore | 35.5 (4.4) |

### Feasibility of Longitudinal Patient Video Uploads

#### Engagement

After enrollment, 44/50 (88%) Discovery cohort participants provided at least 1 time point of video uploads, 2 withdrew from study due to relapses and 4 were lost to follow up.

#### Study Adherence

37/50 (74%) participants uploaded ≥3 weekly assessments and completed the 6 month visit; 2 provided an initial weekly assessment but withdrew due to time demands, and 5 completed >3 weekly assessments but not the 6 month clinical visit. No participants providing remote data experienced relapses. The 44 remotely engaged participants, and the 37 study completers, did not differ from the entire (N=50) cohort in terms of age, sex, race/ethnicity, disease duration or baseline 9HPT (p-value >0.05 for each variable compared (Figure 2)).

**Figure 2.**
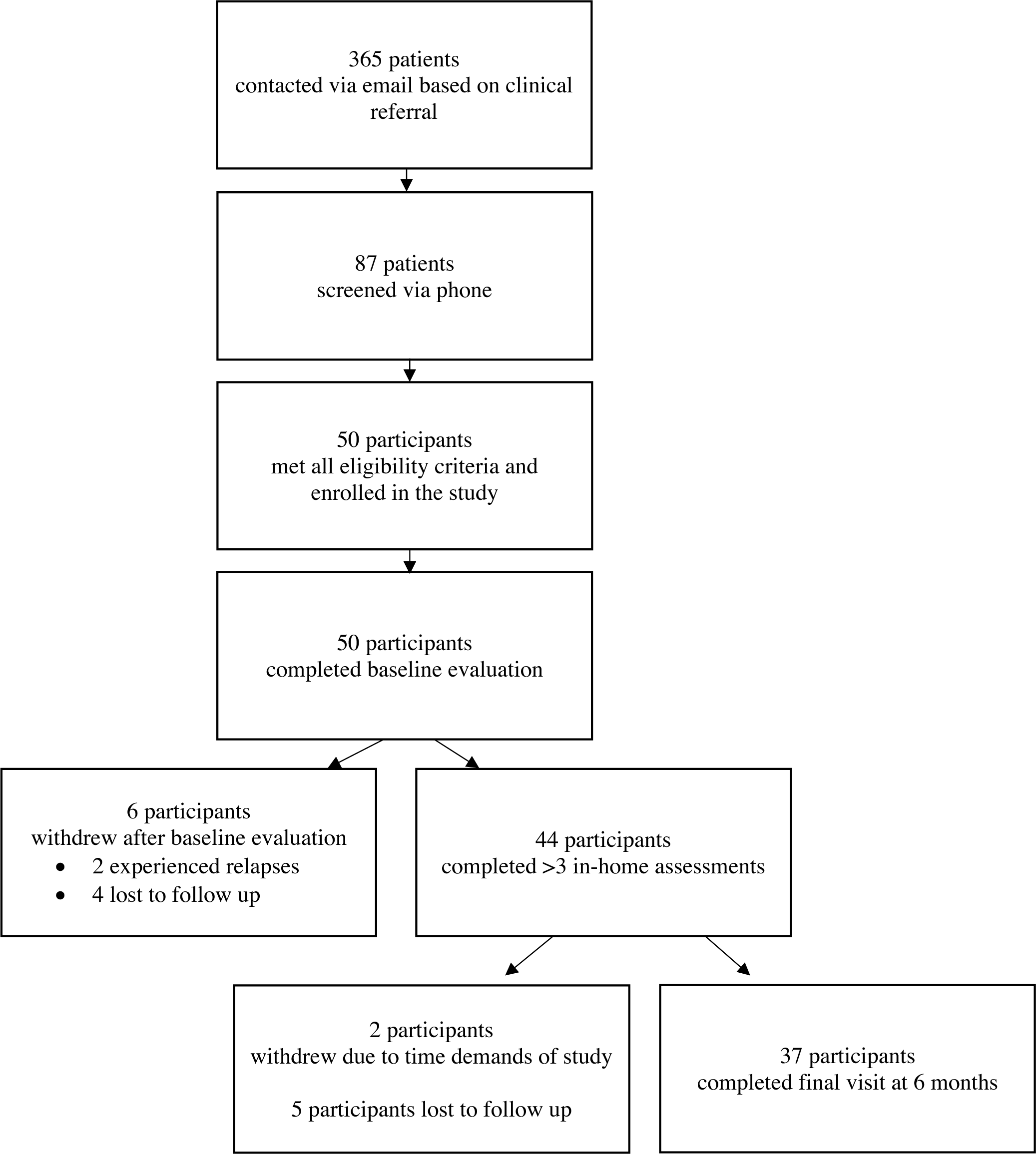
Participant recruitment and retention diagram, tracking patient contacts, enrollments, withdrawals, and completion.

#### Acceptability

The study assessments and overall design were well-received by most participants both in their qualitative and quantitative responses (detailed in Supplementary Table 3).

#### Usability

A majority strongly (77%) or somewhat (18%) agreed that the assessments were easy to access, and a majority strongly (61%) or somewhat (36%) agreed that the remote uploading platform was easy to use, “without technical issues”. None disagreed with these statements.

#### Relevance

Most participants strongly (43%) or somewhat (43%) agreed that the tasks performed in the videos were representative of their daily activities. Qualitatively, buttoning was noted as being “more challenging than I remember”.

### Video quality

Mean video duration was 12.1 seconds (SD 5.6, median 7.2s, range 3-165 seconds). Of 250 total baseline videos (N=50, 5 tasks each), 16 (6.4%) were not analyzed due to low video quality or participant error; of the 234 others, 36 (15.4%) did not generate sufficient data by MediaPipe Hand for kinematic analysis. This left 198 (79%) videos, from 95% participants, to be analyzed. Low (>33.5s)^15^ 9HPT scores were not associated with difficulty using the survey platform or completing video uploads (Pearson’s correlation, p > 0.05).

Longitudinally, the 44 participants with at least 3 complete sets of videos uploaded a mean/median of 11/15 requested timepoints, equivalent to 55/75 requested videos. For the 37 6 month study completers, 345 total videos were uploaded at the baseline and 6 month timepoints; 12 could not be analyzed, and kinematic metrics could not be extracted from 54, leaving 279 (80.9%) videos to be analyzed.

### Cross-sectional validation of video metrics

#### Validation against clinical measures

*Discovery Cohort (N=50).* The buttoning task yielded measures with the strongest correlations with standard in-clinic assessments of hand function (Figure 3). In the non-dominant hand, 9HPT was strongly correlated with wrist position path length (r=0.66, p=0.011), wrist position peaks (r= 0.65, p=0.010), index position path length (r=0.69, p=0.012, and index position peaks (r=0.68, p=0.009). Further, vibration sense was also correlated with wrist position path length (r= 0.39, p=0.045), index position path length (r=0.46, p=0.006), and wrist position peaks (r=0.35, p=0.019). In the dominant hand, these correlations between buttoning metrics and 9HPT ranged r=0.51-0.57, p=0.03-0.05. More modest correlations were found for the brushing and eating tasks (Supplementary Table 6).

**Figure 3.**
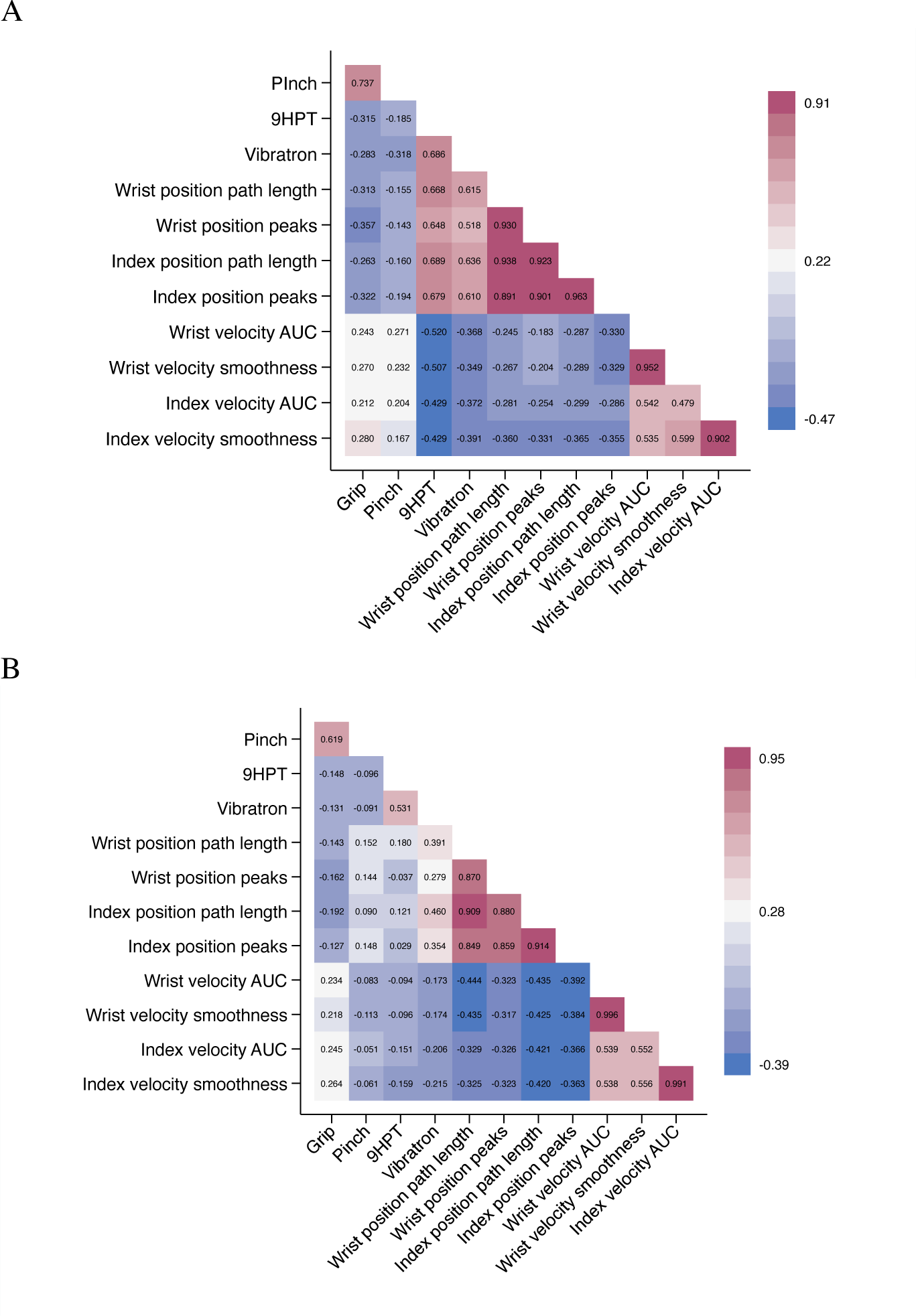
Spearman’s correlation coefficients heatmap, buttoning task. Darker pink colors indicate r values closer to 1, and darker blue colors indicate values closer to −1. (A) Buttoning dominant hand (B) buttoning nondominant hand.

**Figure 4.**
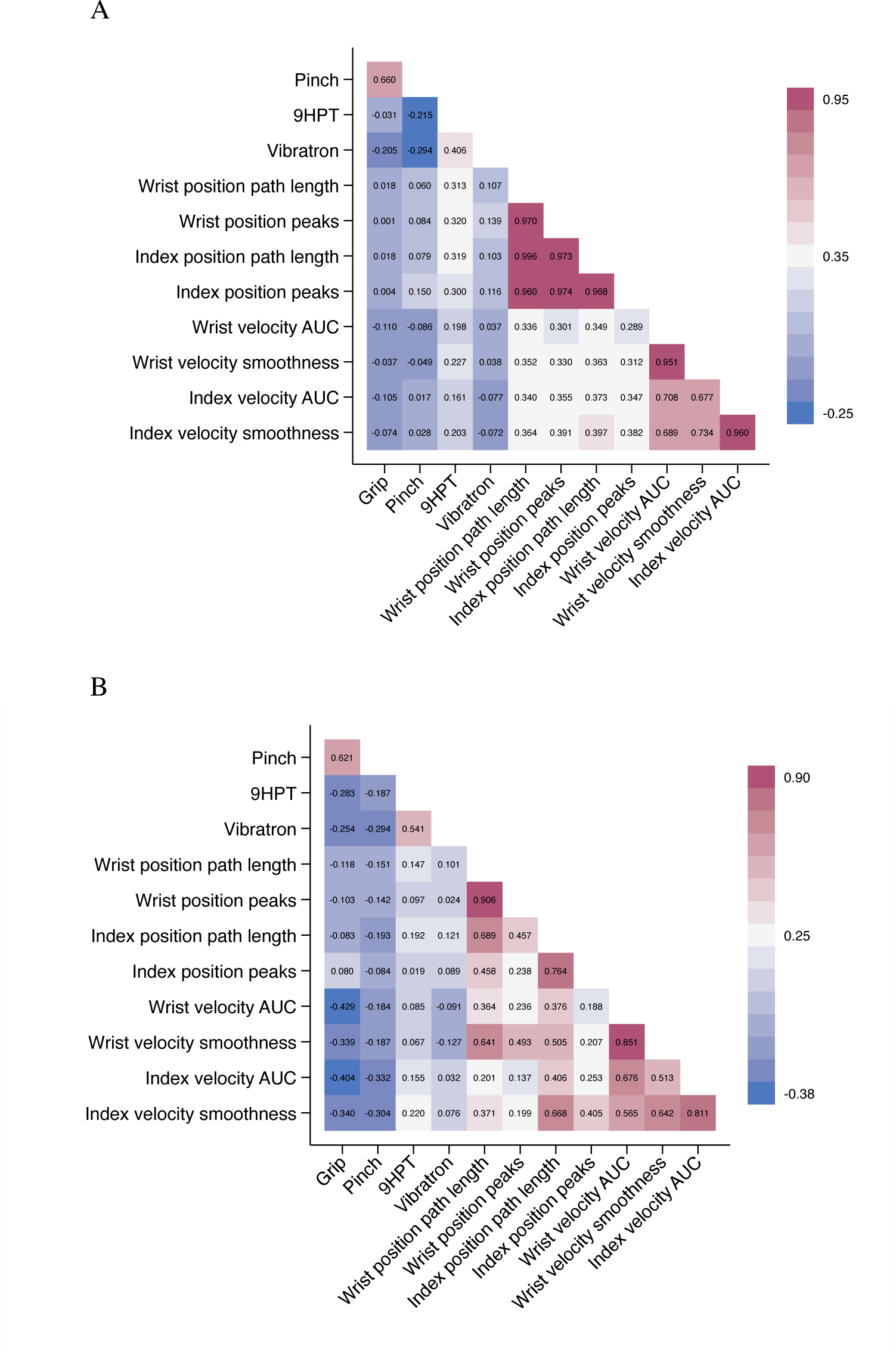
Spearman’s correlation coefficients heatmap, brushing task. Darker pink colors indicate r values closer to 1, and darker blue colors indicate values closer to −1. (A) Brushing dominant hand (B) brushing nondominant hand.

To ensure that these associations were stable across different time points, cross-sectional correlations at 6 months were also analyzed. Similarly, the buttoning task demonstrated the strongest correlations between 9HPT in the non-dominant hand (wrist position path length (r=0.71, p=0.012), wrist position peaks (r= 0.59, p=0.023), index position path length (r=0.61, p=0.019), and index position peaks (r=0.52, p=0.031).

*Validation Cohort (N=35)*. The validation cohort did not differ from the Discovery cohort in any key clinical or demographic features (Supplementary Table 2). Mean 9HPT for the dominant hand was 25.3 seconds and for the nondominant hand was 26.9 seconds. Again, as for the Discovery cohort, the buttoning task yielded measures with the strongest correlations with 9HPT scores. In the non-dominant hand, 9HPT was correlated with wrist position path length (r=0.45, p=0.021) and wrist position peaks (r= 0.42, p=0.010).

#### Video correlates of poor motor control

Greater path of wrist movement while buttoning in the nondominant hand was correlated with both measures poor motor control (difficulty with purposeful, coordinated movements^25^), i.e., long 9HPT time and low pinch strength, as well as diminished vibration sense (high threshold of detection) (Figure 5B). Motor control showed moderate correlations with video metrics from the eating and brushing tasks.

**Figure 5.**
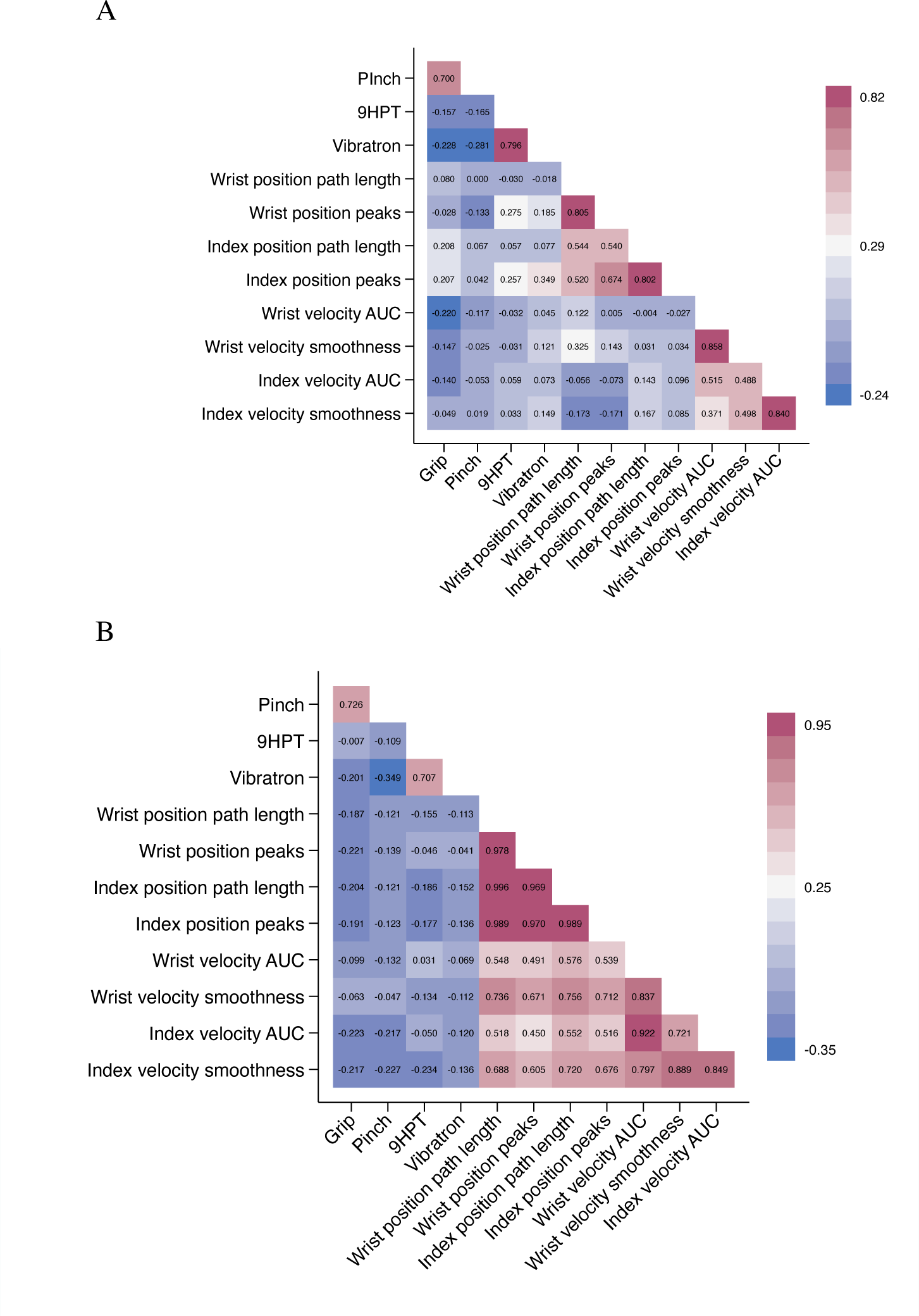
Spearman’s correlation coefficients heatmap, eating task. Darker pink colors indicate r values closer to 1, and darker blue colors indicate values closer to −1. (A) Eating dominant hand (B) eating nondominant hand.

#### Association with patient-perceived dysfunction

Self-reported difficulty in pinch (e.g., fastening a zipper) and grip strength (e.g., opening a jar) on the ABILHAND questionnaire showed stronger associations with buttoning video measures than with the in-clinic 9HPT. For example, self-reported difficulty with pinch showed no correlation with 9HPT (Spearman r= 0.02, p= 0.91), but moderate correlations with wrist position path length (r=-0.48, p=0.05), wrist velocity area under the curve (r=-0.47, p-0.05), and wrist velocity smoothness (r=-0.48, p=0.05) while buttoning.

#### Test-retest reliability

For the 44 participants providing data on consecutive weeks during the first month of the study, metrics for all tasks resulted in r > 0.8, indicating good stability between repeated measures within one week.

### Longitudinal validation of video metrics over 6 months (N=37)

#### Clinical measures

Over the 6 month study, mean 9HPT only decreased by 2.7 seconds for the dominant hand (p=0.11) and 2.4 seconds for the non-dominant hand (p=0.52); 6 participants (15%) had 9HPT scores that changed ≥20% in the dominant hand, and 4 (10%) who changed in the non-dominant hand.^26^ A marginal reduction in pinch strength was noted (mean change dominant hand: 0.8kg/cm^2^, p=0.05; non-dominant hand: 0.6 kg/cm^2^, p=0.16), as well as in the ABILHAND pinch subscore (p=0.05) but not the grasp subscore. Of the 13 participants whose pinch strength worsened in both hands over 6 months, ABILHAND self-reported pinch subscore worsened for 11. Grip strength and vibration were stable over the study period (Supplementary Table 4).

#### Video measures

Based on the strong correlations with 9HPT and with self-reported impairment in the cross-sectional analyses, the buttoning task was selected for the longitudinal analyses. Most metrics derived from each hand showed statistically significant worsening over the 6 months (Table 2). This includes wrist path position (dominant: p=0.04, non-dominant: p=0.05), wrist position peaks (dominant: p=0.01, non-dominant: p= 0.02), wrist velocity smoothness (dominant: p=0.01), and position peaks of the index finger (dominant: p=0.02, non-dominant: p=0.04).

**Table 2.**
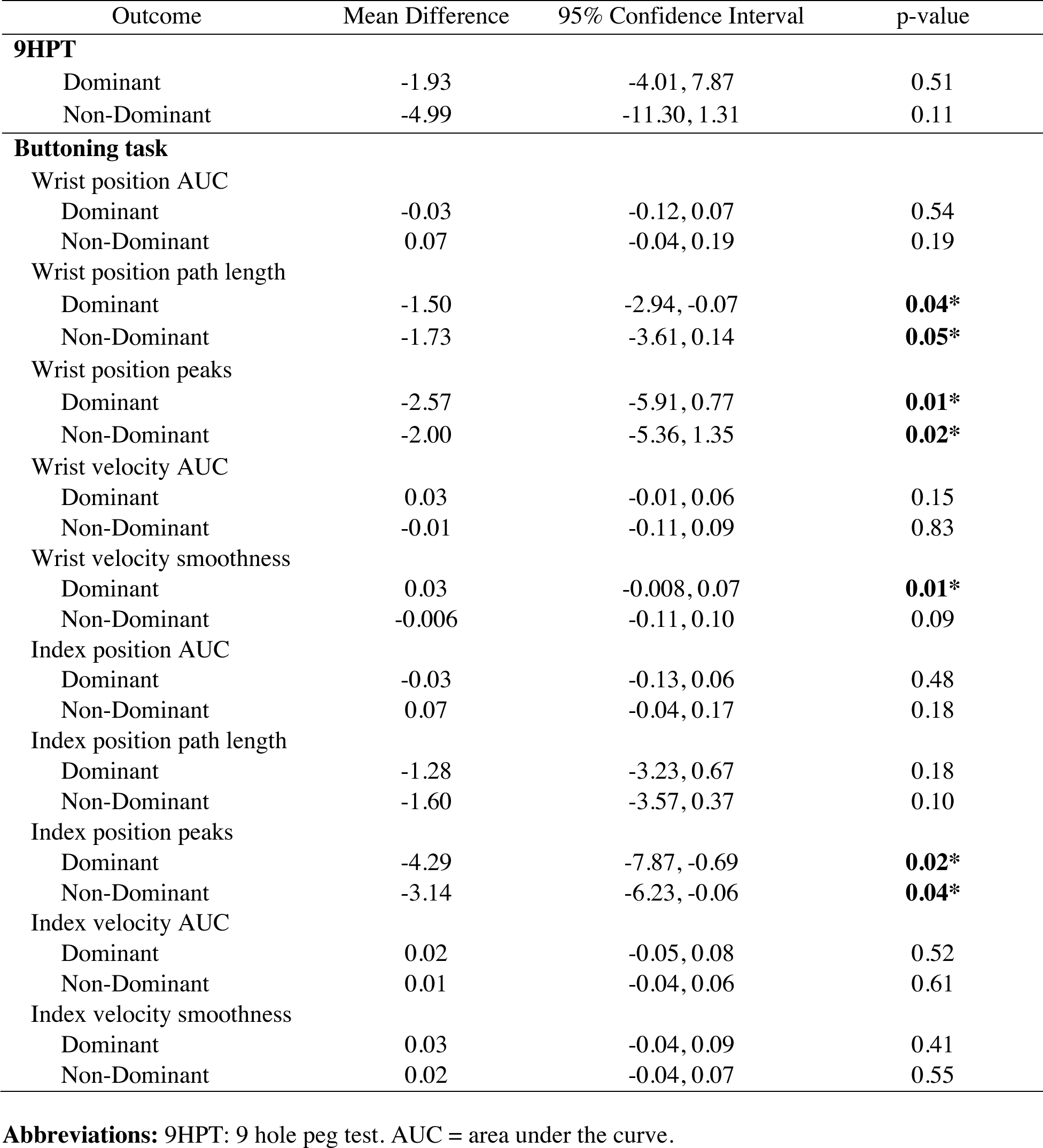
Changes in 9HPT and in video metrics derived from buttoning tasks over 6 months (N=37)

#### Comparative sensitivity to change: clinical vs. buttoning video metrics

Overall, all 37 participants worsened in dexterity, assessed as continuous measures over the study. In the dominant hand, 45% of participants changed by video metrics alone, 35% by both video metrics and 9HPT and 10% by 9HPT alone. The remaining 5% of participants didn’t change on either metric. This trend was similar in the non-dominant hand: 50% of participants changed by video metrics alone, 20% by both video metrics and 9HPT and 15% by 9HPT alone.

When change was measured categorically (i.e., by a 20% worsening), in the dominant hand 60% of participants changed by video metrics alone, 10% by both video metrics and 9HPT and 5% by 9HPT alone. The remaining 25% of participants didn’t change on either metric. Similarly in the non-dominant hand, 65% of participants categorically changed by video metrics alone, 5% by both video metrics and 9HPT, and 5% by 9HPT alone (Figure 6).

**Figure 6.**
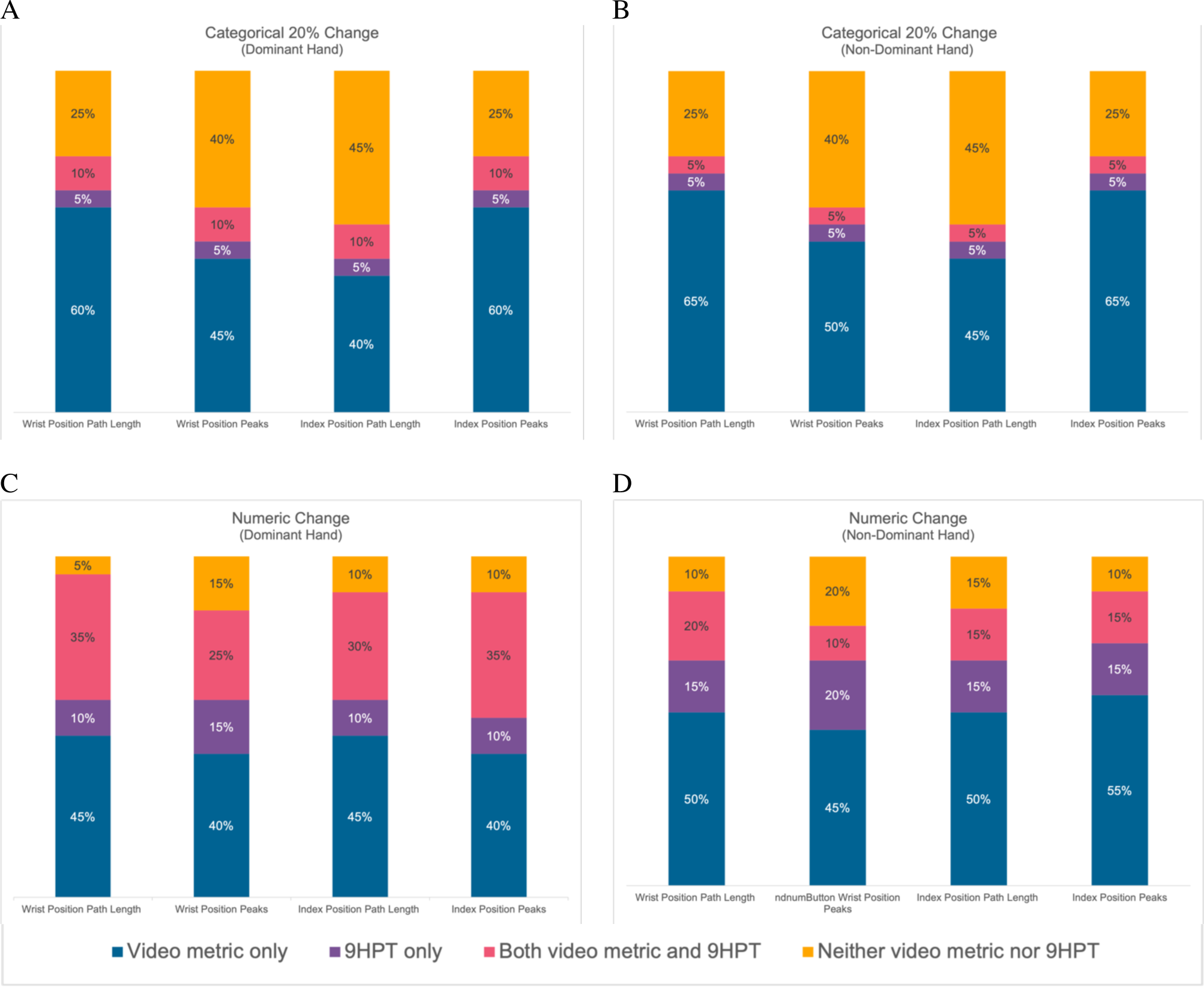
Sensitivity of 9HPT and buttoning video metrics in change detection over study period. Video measures are more sensitive to changes over 6 months than 9HPT. Both categorical 20% change, as well as continuous change, were more commonly detected using video metric than 9HPT scores.

## Discussion

Detecting subtle changes in dexterity has monitoring and treatment implications across many neurological diseases, as well as for the healthy aging population.^27^ The current study sought to overcome limitations of existing in-clinic and remote tools by evaluating a novel form of data collection: patient-generated videos of themselves performing ecologically valid ADLs. In an MS population, this low-cost, low-technology solution was feasible, with high satisfaction and adherence and low barriers to completion.

In proof-of-concept analyses, measures extracted from the buttoning videos correlated highly with the in-clinic 9HPT (and better with self-reported dexterity problems) but were more sensitive to worsening over 6 months despite this being a low disability cohort where only 47% categorically worsened by the established 20% on the 9HPT. The relatively weaker associations and 6-month stability found for brushing and eating tasks are likely due to the larger, gross motor movements required relative to the finer motor abilities required for buttoning and the 9HPT. Buttoning requires the most pinch strength, and given its correlation with vibration sensation, also invokes the importance of dorsal column sensory pathway integrity for manipulating buttons. Further assessments of validity included an independent validation cohort of 35 additional participants with similar 9HPT scores, where again the buttoning task resulted in statistically significant correlations. The stability and replicability of these results speaks to this method’s promise and high clinical validity.

The higher correlations between the patient-reported functional impairments and video measures suggest that they better reflect patient functional capacities than the clinical tests. In fact, most rehabilitation efforts focus on *capacity* measures (what someone reports they are capable of doing, e.g., ABILHAND), though patients seek out rehabilitation services to improve *performance* in their daily, unstructured lives.^28^ Patient-generated videos allow for more direct measurement of movement during tasks in a natural environment, better reflecting performance. Further, identifying the more detailed *aspects* of movement that changed in the video metrics (e.g., speed, distance traveled) could allow clinicians to better understand and target the extent of functional challenges a patient is experiencing.

The current study advances remote research for neurological populations by demonstrating the feasibility and meaningfulness of patient-uploaded videos. Videos provide powerful assessments of movement, and patient-uploaded, in-home videos provide substantial accessibility benefits over videos collected under more controlled in-clinic conditions. Intuitively, uploading “selfies” is near-ubiquitous in modern-day life, and is a technology familiar to many. According to 2019 data, 85% of adult Americans own a smartphone,^29^ including all potential participants referred to the current study (with 44% smartphones being manufactures in 2018 or prior). This “bring your own device” approach to functional assessments overcomes some limitations of other customized digital health approaches (e.g., wearable devices, computerized keyboards, smartphone and tablet-based applications, video conferencing with clinicians), in that it is cost-effective, uses consumer-grade video software both encoded and regularly maintained in most smartphones, and does not require the purchase of hardware. Overall, 95% participants submitted videos that could be analyzed, without any guidance or supervision; and baseline dexterity did not influence their ability to upload high-quality videos and complete assessments. Further, this method delivers videos directly to the clinical or research group who can generate high quality kinematic data without any programming knowledge. The source videos can also be stored for updated analysis as pose estimation algorithms inevitably improve and become more sensitive to impairment and change. The current proof of concept study warrants replication in larger cohorts of individuals with more advanced disability.

The advantages of capturing change through “self-care selfie videos” – including accessibility, adherence and retention, participant satisfaction, correlation with patient-reported difficulties, and sensitivity to change are particularly beneficial given the often insidious, “silent”, disease progression in people with MS. Users’ satisfaction and perceptions of usability are critical to determining the eventual widespread adoption of a digital tool.^30^ Highly specific, easily attainable metrics such as those obtained from the current video analyses could allow clinicians to not only identify limitation in functional activities, but also detect early changes contributing to disease progression and inform rehabilitation protocols. Future applications may include patient-uploaded videos for collection of walking, speech, and other video-amenable data. “Self-care selfies” represent a novel approach to quantify real-time kinematics during real-world ADLs, and this feasible approach could offer benefits across other conditions characterized by loss of hand function (e.g., Parkinson’s Disease, amyotrophic lateral sclerosis) including healthy aging.

## Data Availability

All data produced in the present study are available upon reasonable request to the authors

## Acknowledgement

Dr. Riley Bove is funded by the National Multiple Sclerosis Society Harry Weaver Award. This funding source did not play a role in study design; in the collection, analysis, and interpretation of data; in the writing of the report; nor in the decision to submit the paper for publication.

---

**Supplementary Table 1.** Video Kinematic Metrics Summary.

| **Video Kinematic Metric** | **What is measured?** | **Formula/calculation** |
| --- | --- | --- |
| Position |  |  |
| Path length | How much the joint is moving in 2D space | Sum of the magnitude of the vectors created in 2D space. |
| Local peaks | Movement complexity | MATLAB findpeaks function, which finds peaks from position magnitude curve with a minimum peak prominence of 0.05. |
| Velocity |  |  |
| Area under the curve | Distance traveled by joint | MATLAB function trapz, which approximates integral of velocity magnitude using the trapezoidal method |
| Smoothness | Smoothness of movement | Mean velocity divided by max of non-normalized velocity |

*All video metrics except smoothness are normalized to a zero to one range*

**Supplementary Table 2.** Validation cohort demographics and baseline characteristics.

|  |  |
| --- | --- |
| Gender [N (%)] |  |
| Men | 6 (17%) |
| Women | 28 (80%) |
| Non-binary | 1 (3%) |
| Age [mean (SD)] | 35.0 (15.6) |
| Race [N (%)] |  |
| White, non Hispanic | 25 (71%) |
| Black | 3 (8%) |
| Asian | 6 (17%) |
| Hispanic | 1 (3%) |
| Baseline EDSS [median (IQR)] | 2 (1.5, 3.5) |
| MS Subtype [N (%)] |  |
| Relapsing remitting | 23 (65%) |
| Primary progressive | 5 (14%) |
| Secondary progressive | 6 (17%) |
| Dominant hand [N (%)] |  |
| Right | 30 (85%) |
| Left | 5 (14%) |

**Supplementary Table 3.**
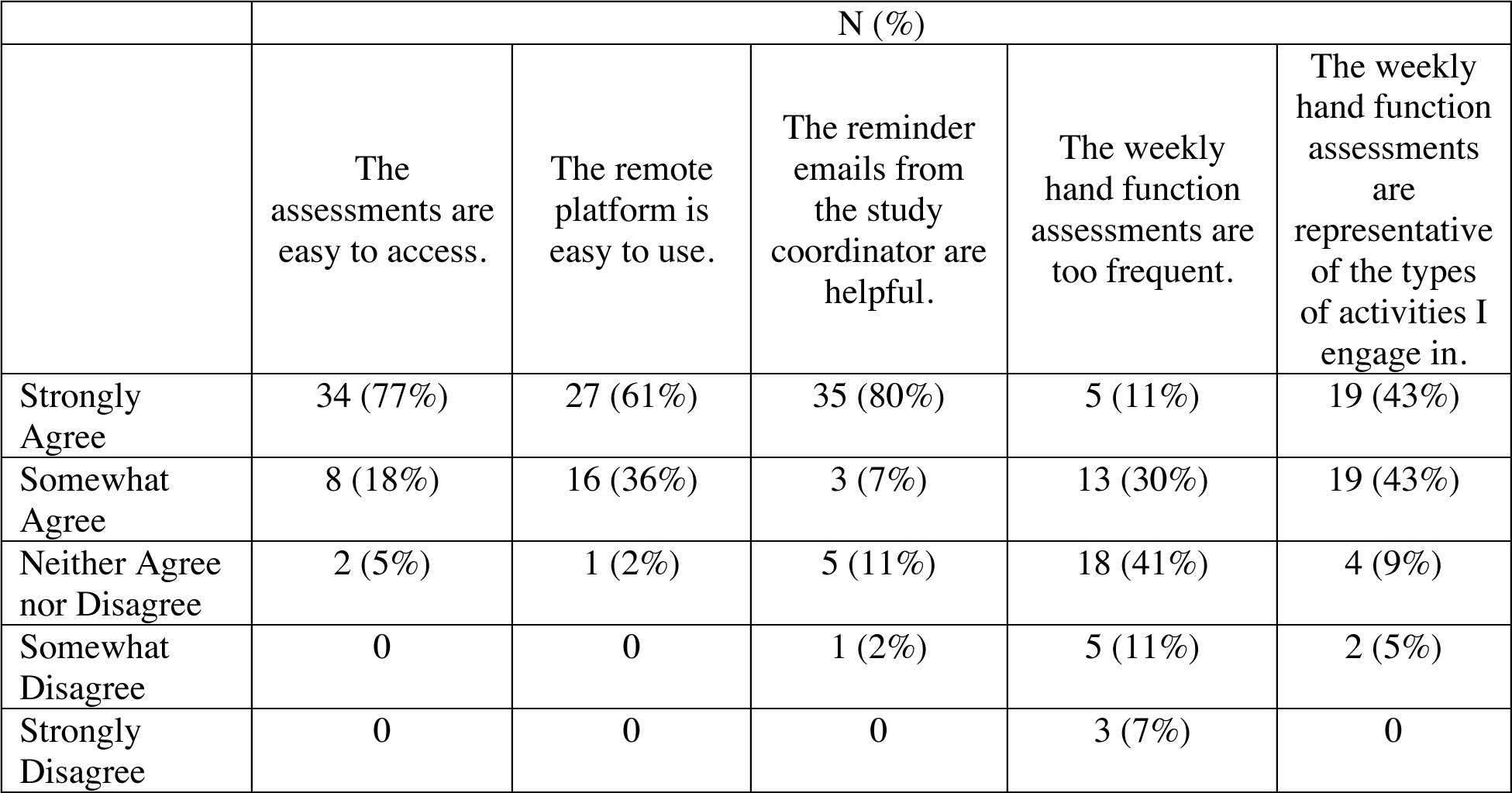
Responses to Monthly Feedback Surveys.

**Supplementary Table 4.** Change in Hand Function Using Established Clinical and Patient-Reported Measures over 6 Months.

| Outcome | Baseline | 6 months | Absolute Change from Baseline to 6 months (N) |  |  | p-value |
| --- | --- | --- | --- | --- | --- | --- |
|  |  |  | Worsened | Improved | No Change |  |
| Participants (N) | 44 | 37 |  |  |  |  |
| **EDSS** |  |  | 10 | 16 | 11 |  |
| Median (IQR) | 3.5 (2, 5) | 2.5 (2, 5) |  |  |  | 0.98 |
| Patients with relapsing MS | 43 | 27 |  |  |  |  |
| **Grip strength (kg/cm<sup>2</sup>)** |  |  |  |  |  |  |
| **[mean (SD)]** |  |  |  |  |  |  |
| Dominant | 28.6 (9.4) | 28.2 (10.5) | 12 | 11 | 14 | 0.84 |
| Non-dominant | 27 (9.5) | 26.6 (10.1) | 13 | 16 | 8 | 0.97 |
| **Pinch strength (kg/cm<sup>2</sup>)** |  |  |  |  |  |  |
| **[mean (SD)]** |  |  |  |  |  |  |
| Dominant | 5.8 (2.6) | 5.0 (2.7) | 13 | 15 | 9 | **0.05\*** |
| Non-dominant | 5.6 (2.5) | 5.0 (2.2) | 13 | 14 | 10 | 0.16 |
| **9HPT (s)** |  |  |  |  |  |  |
| Dominant | 23.5 (9.7) | 20.8 (12.9) | 15 | 9 | 13 | 0.11 |
| Non-dominant | 28.7 (14.9) | 26.3 (18.8) | 13 | 12 | 12 | 0.52 |
| **Vibration sense (Hertz)** |  |  |  |  |  |  |
| **[mean (SD)]** |  |  |  |  |  |  |
| Dominant | 2.4 (2.7) | 3.0 (3.9) | 12 | 11 | 14 | 0.35 |
| Non-dominant | 2.3 (2.6) | 2.5 (3.5) | 12 | 13 | 12 | 0.81 |
| **ABILHAND** |  |  |  |  |  |  |
| Pinch Subscore | 29.5 (4.6) | 29.1 (35.3) | 11 | 9 | 17 | **0.05\*** |
| Grip Subscore | 35.5 (4.4) | 35.3 (4.7) | 7 | 6 | 24 | 0.17 |

**Supplementary Table 6.**
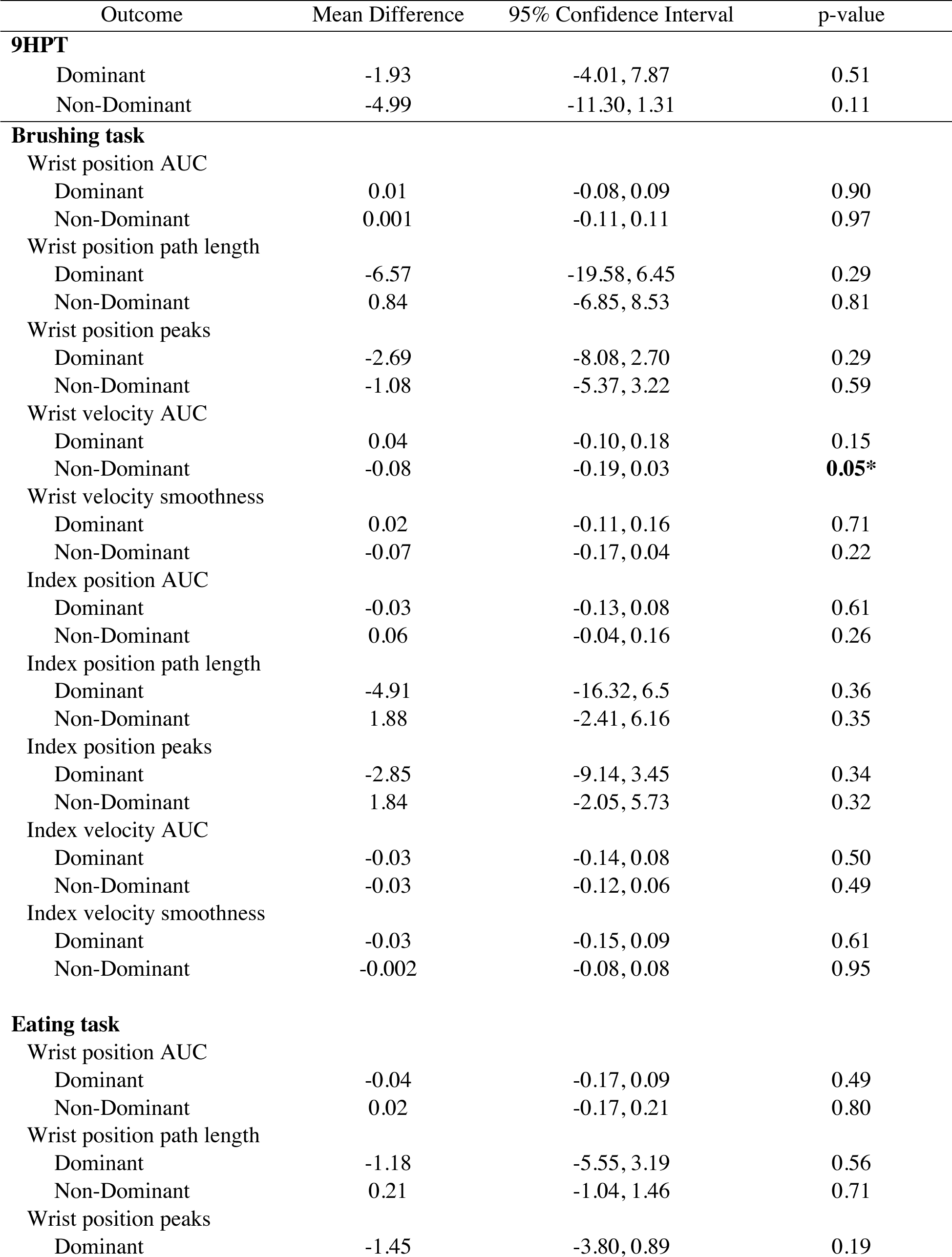
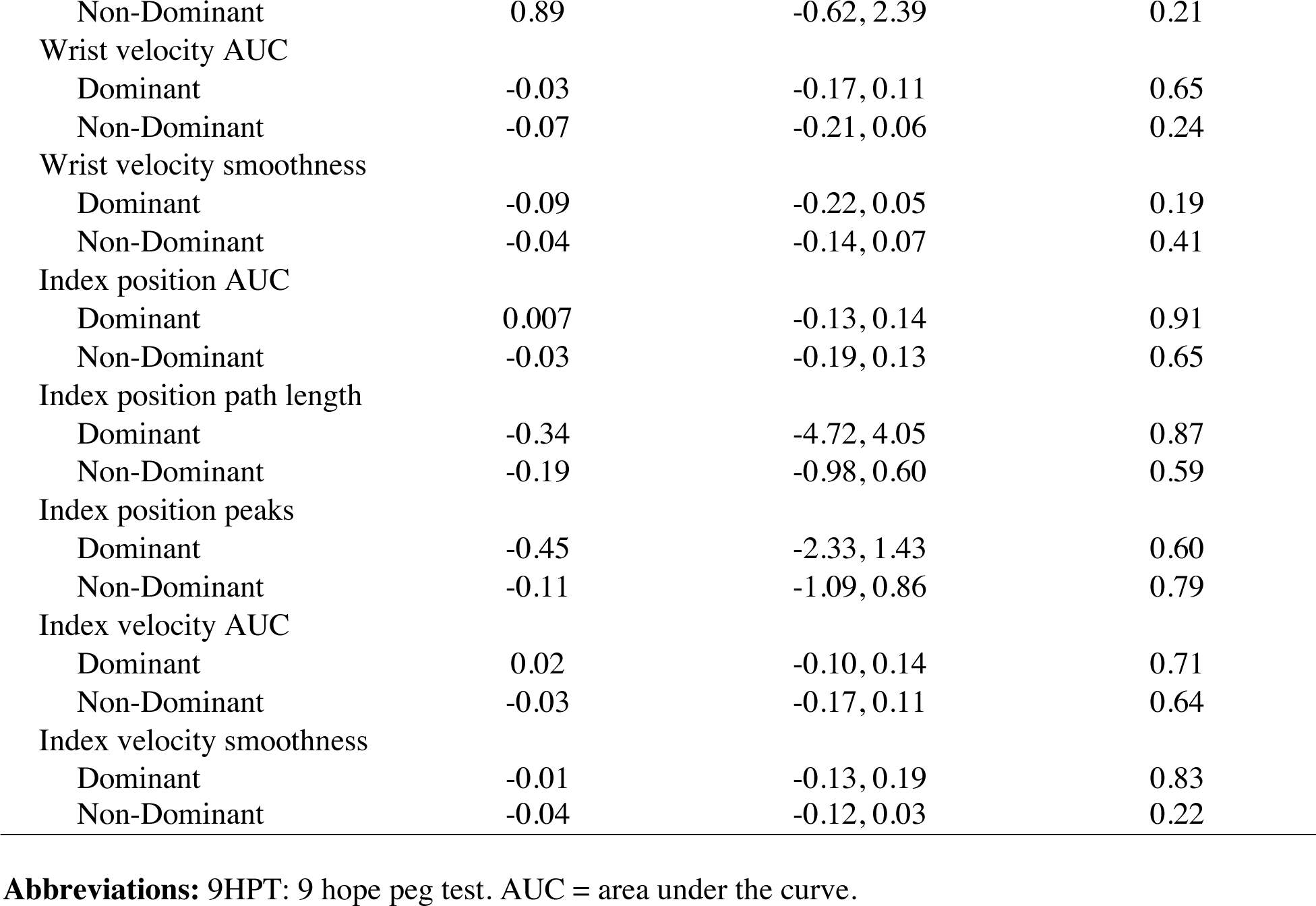
Changes in 9HPT and in video metrics derived from brushing and eating tasks over 6 months (N=37)

| Outcome | Mean Difference | 95% Confidence Interval | p-value |
| --- | --- | --- | --- |
| **9HPT** |  |  |  |
| Dominant | -1.93 | -4.01, 7.87 | 0.51 |
| Non-Dominant | -4.99 | -11.30, 1.31 | 0.11 |
| **Brushing task** |  |  |  |
| Wrist position AUC |  |  |  |
| Dominant | 0.01 | -0.08, 0.09 | 0.90 |
| Non-Dominant | 0.001 | -0.11, 0.11 | 0.97 |
| Wrist position path length |  |  |  |
| Dominant | -6.57 | -19.58, 6.45 | 0.29 |
| Non-Dominant | 0.84 | -6.85, 8.53 | 0.81 |
| Wrist position peaks |  |  |  |
| Dominant | -2.69 | -8.08, 2.70 | 0.29 |
| Non-Dominant | -1.08 | -5.37, 3.22 | 0.59 |
| Wrist velocity AUC |  |  |  |
| Dominant | 0.04 | -0.10, 0.18 | 0.15 |
| Non-Dominant | -0.08 | -0.19, 0.03 | **0.05\*** |
| Wrist velocity smoothness |  |  |  |
| Dominant | 0.02 | -0.11, 0.16 | 0.71 |
| Non-Dominant | -0.07 | -0.17, 0.04 | 0.22 |
| Index position AUC |  |  |  |
| Dominant | -0.03 | -0.13, 0.08 | 0.61 |
| Non-Dominant | 0.06 | -0.04, 0.16 | 0.26 |
| Index position path length |  |  |  |
| Dominant | -4.91 | -16.32, 6.5 | 0.36 |
| Non-Dominant | 1.88 | -2.41, 6.16 | 0.35 |
| Index position peaks |  |  |  |
| Dominant | -2.85 | -9.14, 3.45 | 0.34 |
| Non-Dominant | 1.84 | -2.05, 5.73 | 0.32 |
| Index velocity AUC |  |  |  |
| Dominant | -0.03 | -0.14, 0.08 | 0.50 |
| Non-Dominant | -0.03 | -0.12, 0.06 | 0.49 |
| Index velocity smoothness |  |  |  |
| Dominant | -0.03 | -0.15, 0.09 | 0.61 |
| Non-Dominant | -0.002 | -0.08, 0.08 | 0.95 |
| **Eating task** |  |  |  |
| Wrist position AUC |  |  |  |
| Dominant | -0.04 | -0.17, 0.09 | 0.49 |
| Non-Dominant | 0.02 | -0.17, 0.21 | 0.80 |
| Wrist position path length |  |  |  |
| Dominant | -1.18 | -5.55, 3.19 | 0.56 |
| Non-Dominant | 0.21 | -1.04, 1.46 | 0.71 |
| Wrist position peaks |  |  |  |
| Dominant | -1.45 | -3.80, 0.89 | 0.19 |
| Non-Dominant | 0.89 | -0.62, 2.39 | 0.21 |
| Wrist velocity AUC |  |  |  |
| Dominant | -0.03 | -0.17, 0.11 | 0.65 |
| Non-Dominant | -0.07 | -0.21, 0.06 | 0.24 |
| Wrist velocity smoothness |  |  |  |
| Dominant | -0.09 | -0.22, 0.05 | 0.19 |
| Non-Dominant | -0.04 | -0.14, 0.07 | 0.41 |
| Index position AUC |  |  |  |
| Dominant | 0.007 | -0.13, 0.14 | 0.91 |
| Non-Dominant | -0.03 | -0.19, 0.13 | 0.65 |
| Index position path length |  |  |  |
| Dominant | -0.34 | -4.72, 4.05 | 0.87 |
| Non-Dominant | -0.19 | -0.98, 0.60 | 0.59 |
| Index position peaks |  |  |  |
| Dominant | -0.45 | -2.33, 1.43 | 0.60 |
| Non-Dominant | -0.11 | -1.09, 0.86 | 0.79 |
| Index velocity AUC |  |  |  |
| Dominant | 0.02 | -0.10, 0.14 | 0.71 |
| Non-Dominant | -0.03 | -0.17, 0.11 | 0.64 |
| Index velocity smoothness |  |  |  |
| Dominant | -0.01 | -0.13, 0.19 | 0.83 |
| Non-Dominant | -0.04 | -0.12, 0.03 | 0.22 |

**Abbreviations:** 9HPT: 9 hope peg test. AUC = area under the curve.

## References

1. Smith D. Grasping the Importance of Our Hands. In. InMotion. Vol 16: Amputee Coalition of America; 2006.

2. Cadden M, Arnett P. Factors Associated with Employment Status in Individuals with Multiple Sclerosis. International Journal of MS Care. 2015;17(6):284–291.

3. Feys P, Romberg A, Ruutiainen J, Ketelaer P. Interference of Upper Limb Tremor on Daily Life Activities in People with Multiple Sclerosis. Occupational Therapy In Health Care. 2004;17(3-4):81–95.

4. Fox EJ, Markowitz C, Applebee A, et al. Ocrelizumab reduces progression of upper extremity impairment in patients with primary progressive multiple sclerosis: Findings from the phase III randomized ORATORIO trial. Multiple Sclerosis Journal. 2018;24(14):1862–1870.

5. Aghanavesi S, Nyholm D, Senek M, Bergquist F, Memedi M. A smartphone-based system to quantify dexterity in Parkinson’s disease patients. Informatics in Medicine Unlocked. 2017;9:11–17.

6. Wissel BD, Mitsi G, Dwivedi AK, et al. Tablet-Based Application for Objective Measurement of Motor Fluctuations in Parkinson Disease. Digital Biomarkers. 2017;1(2):126–135.

7. Dai H, Cai G, Lin Z, Wang Z, Ye Q. Validation of Inertial Sensing-Based Wearable Device for Tremor and Bradykinesia Quantification. IEEE Journal of Biomedical and Health Informatics. 2021;25(4):997–1005.

8. Mahadevan N, Demanuele C, Zhang H, et al. Development of digital biomarkers for resting tremor and bradykinesia using a wrist-worn wearable device. npj Digital Medicine. 2020;3(1):5.

9. Akram N, Li H, Ben-Joseph A, et al. Developing and Validating a New Web-Based Tapping Test for Measuring Distal Bradykinesia in Parkinson’s Disease. medRxiv. 2020:2020.2006.2030.20141572.

10. Matarazzo M, Arroyo-Gallego T, Montero P, et al. Remote Monitoring of Treatment Response in Parkinson’s Disease: The Habit of Typing on a Computer. Movement Disorders. 2019;34(10):1488–1495.

11. Kidziński Ł, Yang B, Hicks JL, Rajagopal A, Delp SL, Schwartz MH. Deep neural networks enable quantitative movement analysis using single-camera videos. Nat Commun. 2020;11(1):4054.

12. Sato K, Nagashima Y, Mano T, Iwata A, Toda T. Quantifying normal and parkinsonian gait features from home movies: Practical application of a deep learning-based 2D pose estimator. PLoS One. 2019;14(11):e0223549.

13. Bae JH, Kang SH, Seo KM, Kim DK, Shin HI, Shin HE. Relationship Between Grip and Pinch Strength and Activities of Daily Living in Stroke Patients. Ann Rehabil Med. 2015;39(5):752–762.

14. Zackowski KM, Wang JI, McGready J, Calabresi PA, Newsome SD. Quantitative sensory and motor measures detect change over time and correlate with walking speed in individuals with multiple sclerosis. Multiple Sclerosis and Related Disorders. 2015;4(1):67–74.

15. Feys P, Lamers I, Francis G, et al. The Nine-Hole Peg Test as a manual dexterity performance measure for multiple sclerosis. Mult Scler. 2017;23(5):711–720.

16. Harris PA, Taylor R, Thielke R, Payne J, Gonzalez N, Conde JG. Research electronic data capture (REDCap)--a metadata-driven methodology and workflow process for providing translational research informatics support. J Biomed Inform. 2009;42(2):377–381.

17. Barrett LE, Cano SJ, Zajicek JP, Hobart JC. Can the ABILHAND handle manual ability in MS? Mult Scler. 2013;19(6):806–815.

18. Edemekong PF, Bomgaars DL, Sukumaran S, Levy SB. Activities of Daily Living. In: StatPearls. Treasure Island (FL): StatPearls Publishing StatPearls Publishing LLC.; 2021.

19. Schnall R, Cho H, Liu J. Health Information Technology Usability Evaluation Scale (Health-ITUES) for Usability Assessment of Mobile Health Technology: Validation Study. JMIR Mhealth Uhealth. 2018;6(1):e4.

20. Gionfrida L, Rusli WMR, Bharath AA, Kedgley AE. Validation of two-dimensional video-based inference of finger kinematics with pose estimation. PLoS One. 2022;17(11):e0276799.

21. Cornman HL, Stenum J, Roemmich RT. Video-based quantification of human movement frequency using pose estimation: A pilot study. PLoS One. 2021;16(12):e0261450.

22. Puig-Diví A, Escalona-Marfil C, Padullés-Riu JM, Busquets A, Padullés-Chando X, Marcos-Ruiz D. Validity and reliability of the Kinovea program in obtaining angles and distances using coordinates in 4 perspectives. PLOS ONE. 2019;14(6):e0216448.

23. Romeo AR, Rowles WM, Schleimer ES, et al. An electronic, unsupervised patient-reported Expanded Disability Status Scale for multiple sclerosis. Mult Scler. 2020:1352458520968814.

24. Lamers I, Cattaneo D, Chen CC, Bertoni R, Van Wijmeersch B, Feys P. Associations of upper limb disability measures on different levels of the International Classification of Functioning, Disability and Health in people with multiple sclerosis. Phys Ther. 2015;95(1):65–75.

25. Latash ML, Levin MF, Scholz JP, Schöner G. Motor control theories and their applications. Medicina (Kaunas). 2010;46(6):382–392.

26. Koch MW, Repovic P, Mostert J, et al. The nine hole peg test as an outcome measure in progressive MS trials. Mult Scler Relat Disord. 2023;69:104433.

27. García-Hermoso A, Cavero-Redondo I, Ramírez-Vélez R, et al. Muscular Strength as a Predictor of All-Cause Mortality in an Apparently Healthy Population: A Systematic Review and Meta-Analysis of Data From Approximately 2 Million Men and Women. Archives of Physical Medicine and Rehabilitation. 2018;99(10):2100–2113.e2105.

28. Lang CE, Holleran CL, Strube MJ, et al. Improvement in the Capacity for Activity Versus Improvement in Performance of Activity in Daily Life During Outpatient Rehabilitation. Journal of Neurologic Physical Therapy. 2023;47(1).

29. Technology PRCIa. Mobile Fact Sheet. https://www.pewresearch.org/internet/fact-sheet/mobile/. Published 2021. Updated April 7, 2021. Accessed May 13, 2021.

30. Wienert J, Zeeb H. Implementing Health Apps for Digital Public Health – An Implementation Science Approach Adopting the Consolidated Framework for Implementation Research. Frontiers in Public Health. 2021;9.

